# The effects of intermittent or continuous exercise on renal haemodynamics during moderate-intensity exercise

**DOI:** 10.1101/2024.01.23.24301695

**Authors:** Shotaro Kawakami, Tetsuhiko Yasuno, Saki Kawakami, Ai Ito, Kanta Fujimi, Takuro Matsuda, Shihoko Nakashima, Kosuke Masutani, Yoshinari Uehara, Yasuki Higaki, Ryoma Michishita

## Abstract

**Purpose:** Moderate-intensity exercise has beneficial effects for individuals with CKD. However, it is unclear whether intermittent exercise (IE) has a different effect on renal haemodynamics compared to continuous exercise (CE). This study aimed to compare the effects of intermittent or continuous exercise on renal haemodynamics and renal injury during moderate-intensity exercise.

**Methods:** Ten males underwent IE or CE to consider the effect of exercise on renal haemodynamics during moderate-intensity exercise. Renal haemodynamic assessment and blood-sampling were conducted before exercise (pre) and immediately (post 0), 30-min (post 30), and 60-min (post 60) after exercise. Urine-sampling was conducted in pre, post 0 and post 60.

**Results:** There was no condition-by-time interaction (p = 0.073), condition (p = 0.696), or time (p = 0.433) effects regarding renal blood flow. There was a condition-by-time interaction effect regarding noradrenaline concentrations (p = 0.037). Moreover, both conditions significantly increased noradrenaline concentration at post 0 (IE: p = 0.003, CE: p < 0.001) and remained significantly higher in post 30 (p < 0.001) and post 60 (p < 0.001). Significant difference was found in noradrenaline concentrations at post 0 when comparing IE and CE (399 ± 119 vs. 552 ± 224 pg/ml, p = 0.037). Urinary neutrophil gelatinase-associated lipocalin concentrations increased at post 60 (p = 0.009), but none of them exceeded the cutoff values for the definition of renal damage. Other renal injury biomarkers showed a similar pattern.

**Conclusion:** These findings suggest that IE has a similar effect on renal haemodynamics and function, and AKI biomarkers compared to CE.

## Introduction

Recent reports demonstrate that moderate-intensity continuous exercise (MICE) has beneficial effects for individuals with chronic kidney disease (CKD) (Afsar et al. 2018; Vanden Wyngaert et al. 2018; Watson et al. 2018; Thompson et al. 2019). Accordingly, moderate intensity has been the recommended intensity for patients with CKD and may be quite attractive an option for these patients (Liguori et al. 2021). However, whether MICE affects renal haemodynamics and induces kidney injury remains unclear. Therefore, we recently investigated the influence of MICE on the kidneys and demonstrated that MICE maintains renal blood flow (RBF) and does not increase acute kidney injury (AKI) biomarkers (Kawakami et al. 2022).

Physical inactivity has deleterious effects on CKD progression (Zelle et al. 2017) and is an independent risk factors for CKD development, in addition to aging and obesity (Hallan et al. 2006). In contrast, physical activity, aerobic capacity, and muscle mass decrease even in the early stages of CKD, and continue to decrease with disease progression (Zelle et al. 2017; Kirkman et al. 2021). Accordingly, physical inactivity, decreased aerobic capacity, and reduced renal function cause a vicious cycle that must be broken. Previous studies investigated the associations between physical activity and the incidence and prevalence of CKD and revealed that regular exercise and higher physical activity suppress renal function decline (Michishita et al. 2016a, b, 2017). Therefore, regular exercise and increased physical activity may be key factors in the suppression of dialysis introduction and extension of healthy life expectancy (Beetham et al. 2022). However, patients with CKD patients with lower cardiorespiratory fitness and muscle strength (Kirkman et al. 2021) are unable to perform continuous exercise, requiring the exploration of practicable exercise conditions. For some individuals with CKD who are unable to perform continuous exercise, the American College of Sports Medicine’s (ACSM) guidelines state that these patients should perform moderate-intensity exercise at intervals (Liguori et al. 2021). A previous study of the acute effects of moderate-intensity intermittent exercise (MIIE) and MICE on arterial stiffness demonstrated that MIIE elicited a greater decrease in arterial stiffness than MICE (Zheng et al. 2015). In addition, a recent report (Meric et al. 2022) described a statistically significantly lower blood lactate concentration observed after MIIE than after MICE, suggesting that MIIE may be less metabolically effective in causing fatigue. However, to the best of our knowledge, no studies have examined the effects of MIIE on renal haemodynamics and function. Therefore, exploring the influence of MIIE on renal haemodynamics and function is important to provide practicable exercise conditions.

The hypothesis of this study is that MIIE has a more favourable effect on renal haemodynamics and function compared to MICE. To test this hypothesis, we compared the effects of MIIE and MICE on renal haemodynamics and function.

## Material and Methods

### Participants

This study included 10 healthy adult males (age: 37 ± 8 yr; height: 1.72 ± 0.05 m; weight: 67.4 ± 6.0 kg) (Table 1). The inclusion and exclusion criteria were as follows: the presence or absence of 1) estimated glomerular filtration rate (eGFR) ≥ 60 ml/min/1.73 m^2^, 2) serious or progressive disease and symptoms, 3) any specific underlying medical conditions (cardiovascular or cerebrovascular disease, receiving dialysis) or a history of associated symptoms, and 4) taking medications that affect systemic haemodynamics. At the time of designing the present study, to our knowledge, there were no published data on the effect size of MIIE manipulation on renal haemodynamics. Therefore, a power calculation performed in a prior study that investigated the effect of MIIE on endothelial function indicated that 12 participants would be estimated to be statistically appropriate, with a statistical power of 0.8 and an alpha of 0.05 (Meric et al. 2022). Participants underwent a medical checkup, including an electrocardiogram, prior to participation to ensure that they were appropriate for this study. Twelve healthy, middle-aged adults were recruited in this study. However, one participant did not complete the experiment because of an unrelated injury, and one participant was unable to complete the experiment because of participant availability. Consequently, the data are presented as n = 10 throughout the experiment. Participants avoided strenuous exercise the day before testing, fasted 8 h prior to testing (drinking water was acceptable), and avoided breakfast, caffeine, and exercise on the test day. Although the MICE condition included data from previous study (Kawakami et al. 2024), the purpose of that study was to compare the effects of high-intensity intermittent exercise and MICE on renal haemodynamic parameters, whereas the purpose of this study was the effects of continuous or intermittent exercise on renal haemodynamics during moderate-intensity exercise. Therefore, this study excluded duplicate publication.

**Table 1.** Participant characteristics.

|  |  |
| --- | --- |
| Age, yr | 37 ± 8 |
| Height, m | 1.72 ± 0.05 |
| Weight, kg | 67.4 ± 6.0 |
| Body Mass Index, kg/m<sup>2</sup> | 22.9 ± 2.0 |
| Systolic blood pressure, mmHg | 110 ± 7 |
| Diastolic blood pressure, mmHg | 74 ± 9 |
| Serum creatinine, mg/dL | 0.86 ± 0.07 |
| eGFR, ml/min/1.73m<sup>2</sup> | 82 ± 9 |
| VO<sub>2</sub>peak, ml/kg/min | 37.0 ± 9.9 |
| VO<sub>2</sub>@LT, ml/kg/min | 13.2 ± 4.6 |

Data are expressed as the mean ± SD.
**eGFR** estimated glomerular filtration rate, **LT** lactate threshold, **VO<sub>2</sub>peak** peak oxygen uptake

### Incremental exercise test for determination of optimal exercise intensity

All participants performed an incremental exercise test using a cycle ergometer (Lode; Corival, the Netherlands) to determine optimal exercise intensity as described previously (Kawakami et al. 2018). Blood was drawn from the ear lobe at rest and at 1-min intervals during exercise to determine blood lactate concentrations. In this study, we defined the lactate threshold (LT) intensity that exhibited no decrease in RBF as moderate-intensity based on our previous findings (Kawakami et al. 2018). The LT was decided by five technicians, and each technician assessed the steep-increase point of the LA by visual inspection of graphical plots of LA versus workload. LT was defined as the means of three of five values, excluding the maximum and the minimum values obtained from the participants.

### The consideration of the impacts of MICE and MIIE on renal haemodynamics

On arrival, each participant provided urine samples, drank water, and rested for 5 minutes. Subsequently, renal haemodynamics were assessed using ultrasound echo. Blood samples were collected to determine the blood biochemistry following a renal haemodynamic assessment. Each participant performed MIIE (5-min at LT intensity and 1-min at rest) or MICE (30-min at LT intensity) using a cycle ergometer (Fig. 1). MIIE was performed for 5 min with a 1 min break between them and was repeated six times so that the total energy consumption under both exercise conditions was the same. To match the total energy consumption of the MIIE and MICE conditions, the energy expenditure in both conditions was calculated using the following equation (Liguori et al. 2021): Energy expenditure (kcal) = [[[3.5 + 3.5 + [1.8 × workload (watts) × 6.12]/body weight (kg)]/3.5]–1] × exercise duration (hours) × body weight (kg) × 1.05. Renal haemodynamic assessment and blood sampling were performed before exercise (*Pre*), immediately after exercise (*Post 0*), 30 minutes (*Post 30*), and 60 minutes (*Post 60*) after exercise. Urine samples were collected *Pre*, *Post 0*, and *Post 60*. HR and RPE were consistently measured throughout the experiment, and blood pressure was measured *Pre* and *Post 0*. We evaluated the influence of the (dis-) continuity of exercise on renal haemodynamics using ultrasonography. Blood and urine samples were collected to explore the mechanisms responsible for renal haemodynamic regulation and renal injury following exercise. Furthermore, the above two exercise conditions were performed in random order, with at least one week as the washout period between the two conditions. The participants received free hydration during and following the experiment.

**Fig. 1.**
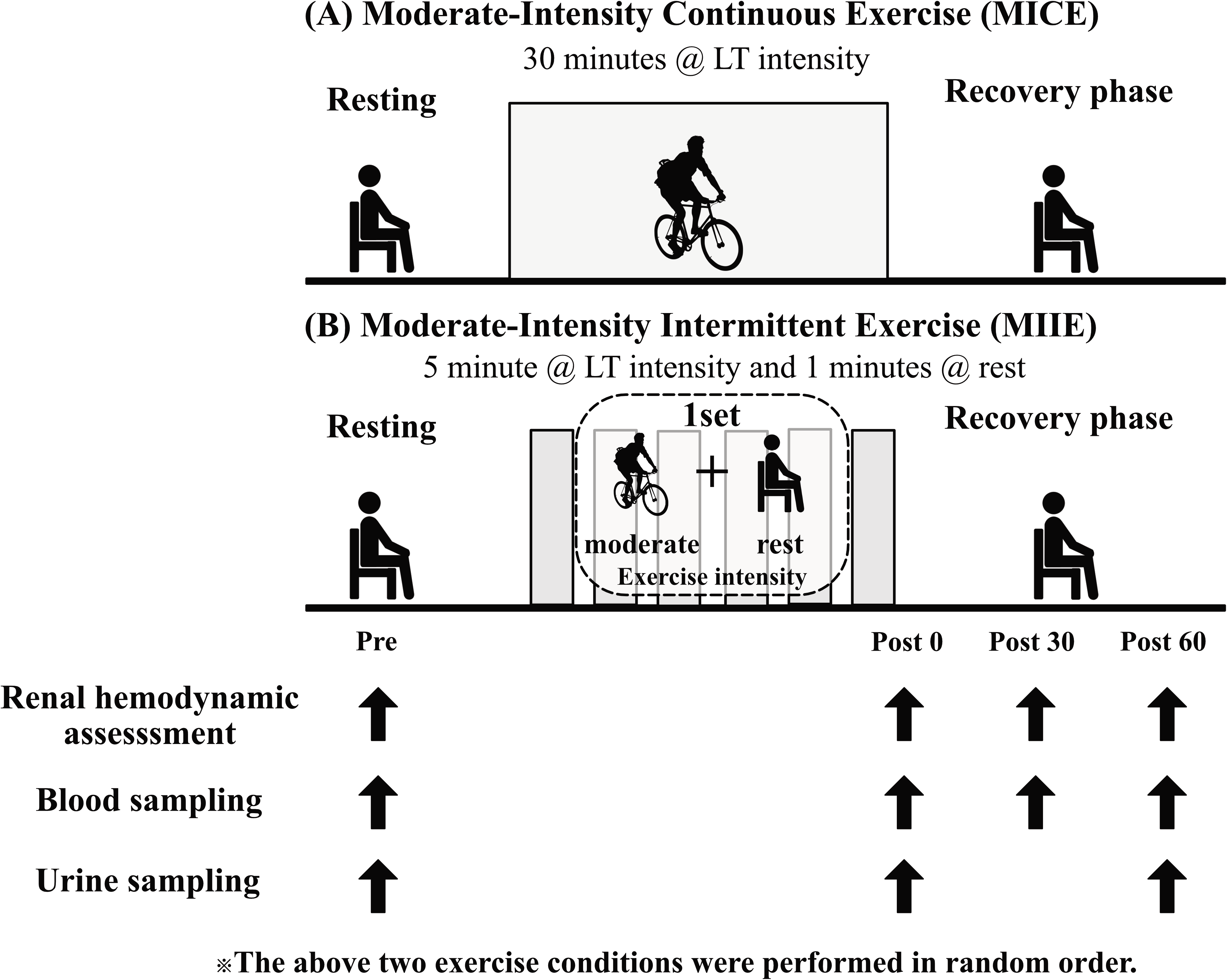
Experimental protocol. All participants performed (**A**) 30 minutes continuous exercise and/or (**B**) intermittent exercise at LT intensity using cycle ergometer. Moderate-intensity intermittent exercise was performed for 5 min with 1 min break between them, and it repeated 6 times so that the total energy consumption of both exercise conditions was the same. The above two exercise conditions were performed in random order, with at least one week as a washout period between the two conditions. Renal haemodynamics assessment and blood-sampling were performed before exercise (*Pre*), immediately after exercise (*Post 0*), 30 minutes (*Post 30*) and 60 minutes (*Post 60*) after exercise. Urine samples were taken in *Pre*, *Post 0* and *Post 60*

### The assessment of renal haemodynamics

Renal haemodynamics were assessed by pulse Doppler using a 3.5 MHz convex electronic scanning probe of an ultrasound system (Aplio 300; Toshiba Medical Systems), as described previously (Kawakami et al. 2024). RBF, blood flow velocity (BFV), and cross-sectional area (CSA) were examined in this study. There are several advantages to evaluate renal haemodynamics using ultrasound echo, which can non-invasively visualise blood vessels by applying a probe, leading to a reduction in the burden on the participants. Another benefit of ultrasound echo is that it evaluates the BFV and CSA of the renal artery in the kidney independently, enabling the examination of whether the variation in RBF is derived from variations in BFV and/or CSA.

### The determination of blood and urinary biomarkers

Blood and urinary biomarkers were determined as described in our previous study (Kawakami et al. 2022). Blood and urine samples were collected from the antecubital vein in the morning after an 8-h overnight fast, and adrenaline, noradrenaline, plasma renin activity (PRA), aldosterone, serum creatinine (sCr), and cystatin C (sCys) levels were measured. In addition, all participants provided urine samples to examine urinary creatinine (uCr), albumin (uAlb), liver-type fatty acid-binding protein (uL-FABP), and N-acetyl-beta-d-glucosaminidase (uNAG) levels to assess renal injury in response to exercise. Furthermore, we measured urinary kidney injury molecule 1 (uKIM-1) in duplicate using a sandwich ELISA kit (Human Urinary KIM-1 Quantikine ELISA Kit, R&D Systems, United States) and urinary neutrophil gelatinase-associated lipocalin (uNGAL) in duplicate using a sandwich ELISA kit (Human NGAL Quantikine ELISA Kit, R&D Systems, United States) to examine kidney injury response to exercise. Each blood sample was centrifuged for 10 min at 1750×*g* at 4 ℃, and a part of the urine sample was centrifuged for 5 min at 400×*g* at 4 ℃. Samples were stored at -80℃ until analysis. The analyses were performed by a commercial blood and urine company (LSI Medience Corp., Tokyo, Japan). Furthermore, eGFR with sCr or sCys and filtration fraction (FF) as indicators of renal function and renal haemodynamics, was calculated as described in our previous study (Kawakami et al. 2022).

### Statistical analysis

The influence of a single bout of MIIE and MICE on all outcome measures was analysed using a two-way analysis of variance. Bonferroni’s *post hoc* test was performed for significant main or interaction effects. All statistical analyses were performed using Prism version 10.0.2 (GraphPad Software, San Diego, CA, USA). P < 0.05 was considered statistically significant. Experimental data are shown as the mean ± standard deviation.

## Results

### Renal haemodynamics following MIIE and MICE conditions

Figure 2 shows the influence of continuity during moderate intensity exercise on renal haemodynamics. There were no condition-by-time interaction, time, or condition effects on the changes in RBF (Fig. 2A), BFV (Fig. 2B) and CSA (Fig. 2C).

**Fig. 2.**
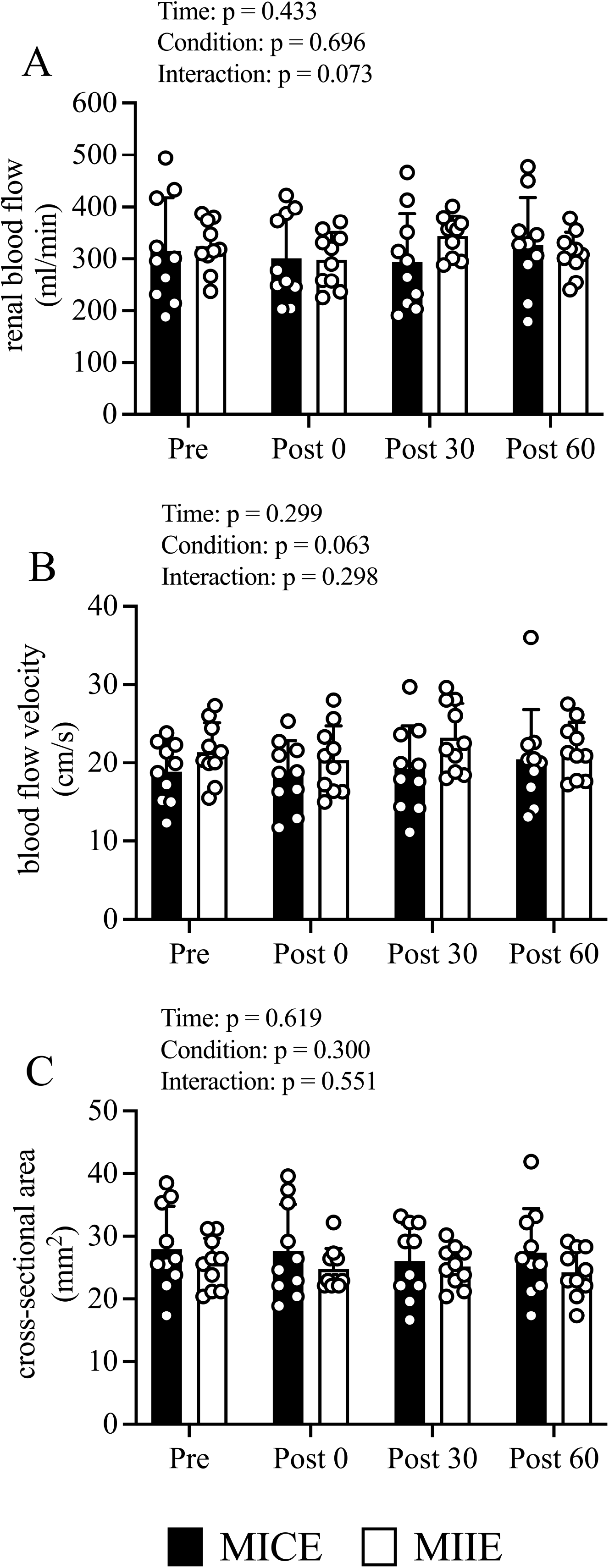
Changes in renal blood flow (**A**), blood flow velocity (**B)** and cross-sectional area (**C)** from before exercise (*Pre*) to immediately after exercise (*Post 0*) to 30 min post-exercise (*Post 30*) to 60 min post-exercise (*Post 60*) in MICE (filled bar, n = 10) and MIIE (open bar, n = 10). Data are the mean ± standard deviation. Open circles represent individual data. Multiple pairwise comparisons were corrected using the Bonferroni method

### Renal function and injury

Figure 3 shows the uCr, uAlb, uNAG, uL-FABP, uKIM-1, and uNGAL excretion responses to MIIE and MICE, all AKI biomarkers that were corrected for uCr levels. There were no condition-by-time interaction or condition effects; however, there was a time effect (p = 0.006) for uCr (Fig. 3A). The uCr exhibited no significant changes at *Post 0* compared with *Pre* and exhibited a significant decrease at *Post 60* (p = 0.003) in both conditions. There were no condition-by-time interaction, time, or condition effects on changes in uAlb (Fig. 3B), uNAG (Fig. 3C), uL-FABP (Fig. 3D) levels. There were no condition-by-time interaction or condition effects; however, there was a time effect for the uKIM-1 (Fig. 3E; p = 0.014) and uNGAL levels (Fig. 3F; p = 0.016). The uKIM-1 levels exhibited no significant changes at *Post 0* compared to *Pre* and exhibited a significant decrease at *Post 60* (p = 0.009) in both conditions. The uNGAL levels had no significant changes at *Post 0* compared to *Pre* and exhibited a significant increase at *Post 60* (p = 0.009) in both conditions.

**Fig. 3.**
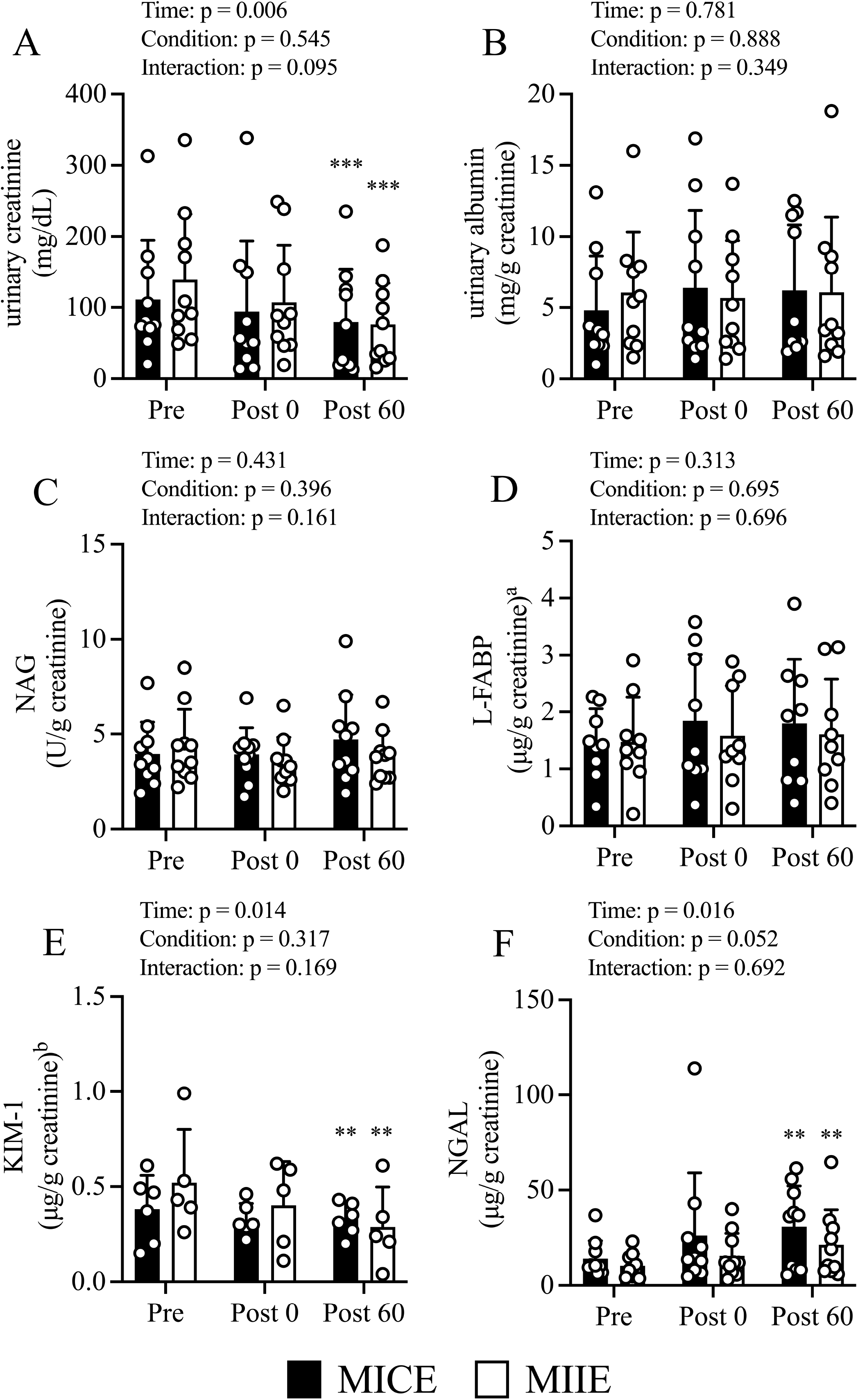
Changes in acute kidney injury biomarkers before and after exercise and recovery phase. Changes in urinary creatinine (**A**), urinary albumin (**B**), N-acetyl-beta-d-glucosaminidase (**C**), liver-type fatty acid-binding protein (**D**), kidney molecule injury 1 (**E**), neutrophil gelatinase-associated lipocalin (**F**) from before exercise (*Pre*) to immediately after exercise (*Post 0*) to 60 min post-exercise (*Post 60*) in MICE (filled bar, n = 10) and MIIE (open bar, n = 10). Data are the mean ± standard deviation. * p < 0.05, ** p < 0.01, *** p < 0.005 vs. *Pre*. ♯ p < 0.05, ♯♯ p < 0.01 vs. MICE. ^a^Data are available as follows: MICE, n = 9; MIIE, n = 9. ^b^Data are available as follows: MICE, n = 6; MIIE, n = 5. Open circles represent individual data. Multiple pairwise comparisons were corrected using the Bonferroni method

Moreover, there were no condition-by-time interaction, time, or condition effects on the changes in sCr (Fig. 4A), sCys (Fig. 4B), eGFR_cre_ (Fig. 4C), eGFR_cys_ (Fig. 4D), FF_cre_ (Fig. 4E), or FF_cys_ (Fig. 4F) levels.

**Fig. 4.**
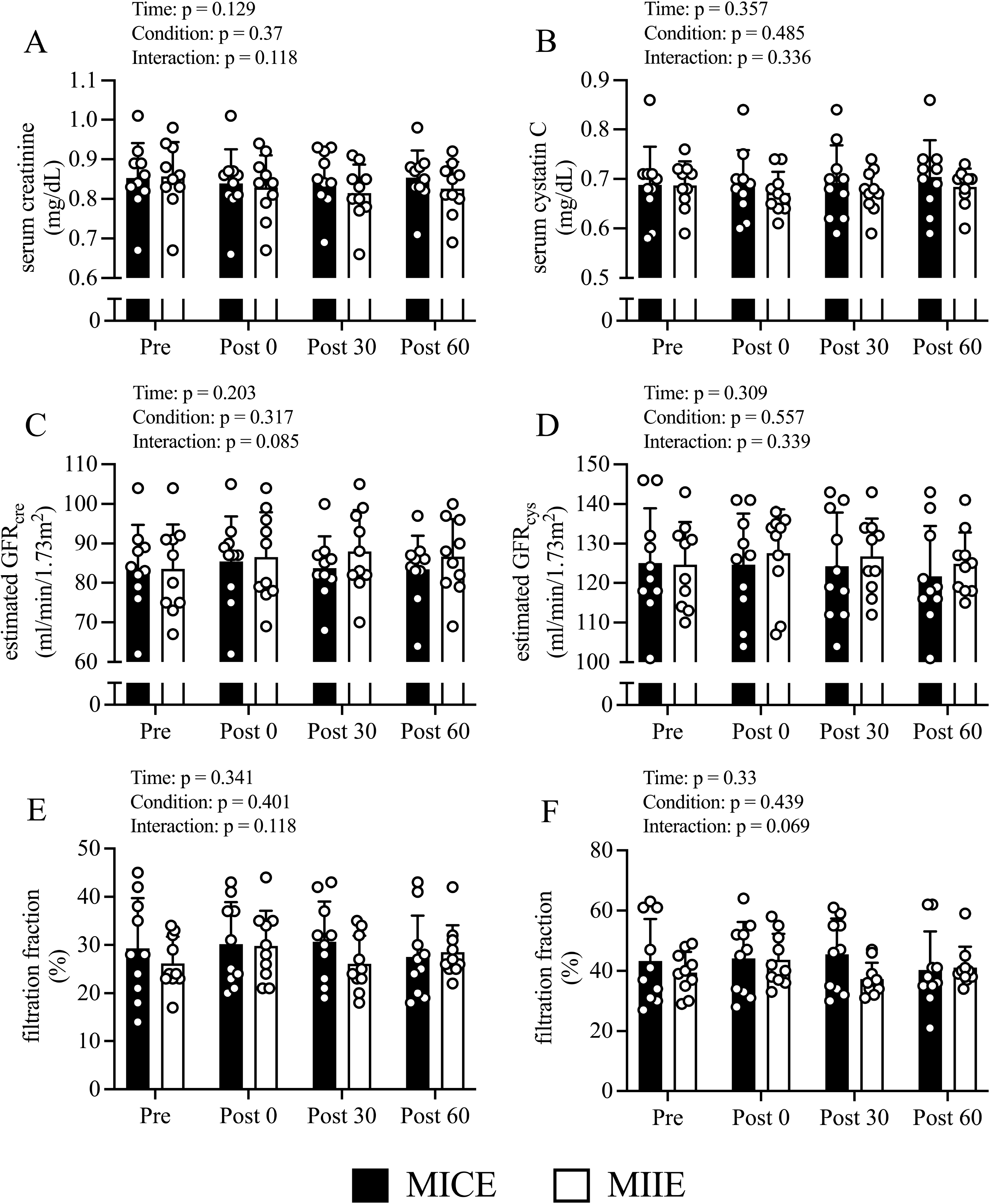
Changes in renal function parameters before and after exercise and recovery phase. Changes in serum creatinine (**A**), serum cystatin C (**B**), estimated GFR_cre_ (**C**), estimated GFR_cys_ (**D**), filtration fraction (creatinine) (**E**), filtration fraction (cystatin C) (**F**) from before exercise (*Pre*) to immediately after exercise (*Post 0*) to 30 min post-exercise (*Post 30*) to 60 min post-exercise (*Post 60*) in MICE (filled bar, n = 10) and MIIE (open bar, n = 10). Data are the mean ± standard deviation. ***GFR*** glomerular filtration rate. Open circles represent individual data. Multiple pairwise comparisons were corrected using the Bonferroni method

### Biochemical parameters associated with the regulation of renal haemodynamics

Changes in catecholamines and measures of the renin-angiotensin system (RAS) before and after the exercise and recovery phase are shown in Figure 5. A condition-by-time interaction effect was observed for noradrenaline (Fig. 5A; p = 0.037). Noradrenaline at Pre did not differ between the conditions; however, the noradrenaline level at Post 0 was lower in the MIIE condition than in the MICE condition (p = 0.037). In the MIIE condition, noradrenaline exhibited a significant increase at Post 0 compared to Pre (p = 0.003) and remained significantly higher at Post 30 (p < 0.001) and Post 60 (p < 0.001). In contrast, noradrenaline showed a significant increase at Post 0 compared to Pre (p < 0.001) and remained significantly higher at Post 30 (p < 0.001) and Post 60 (p = 0.001) in the MICE condition.

**Fig. 5.**
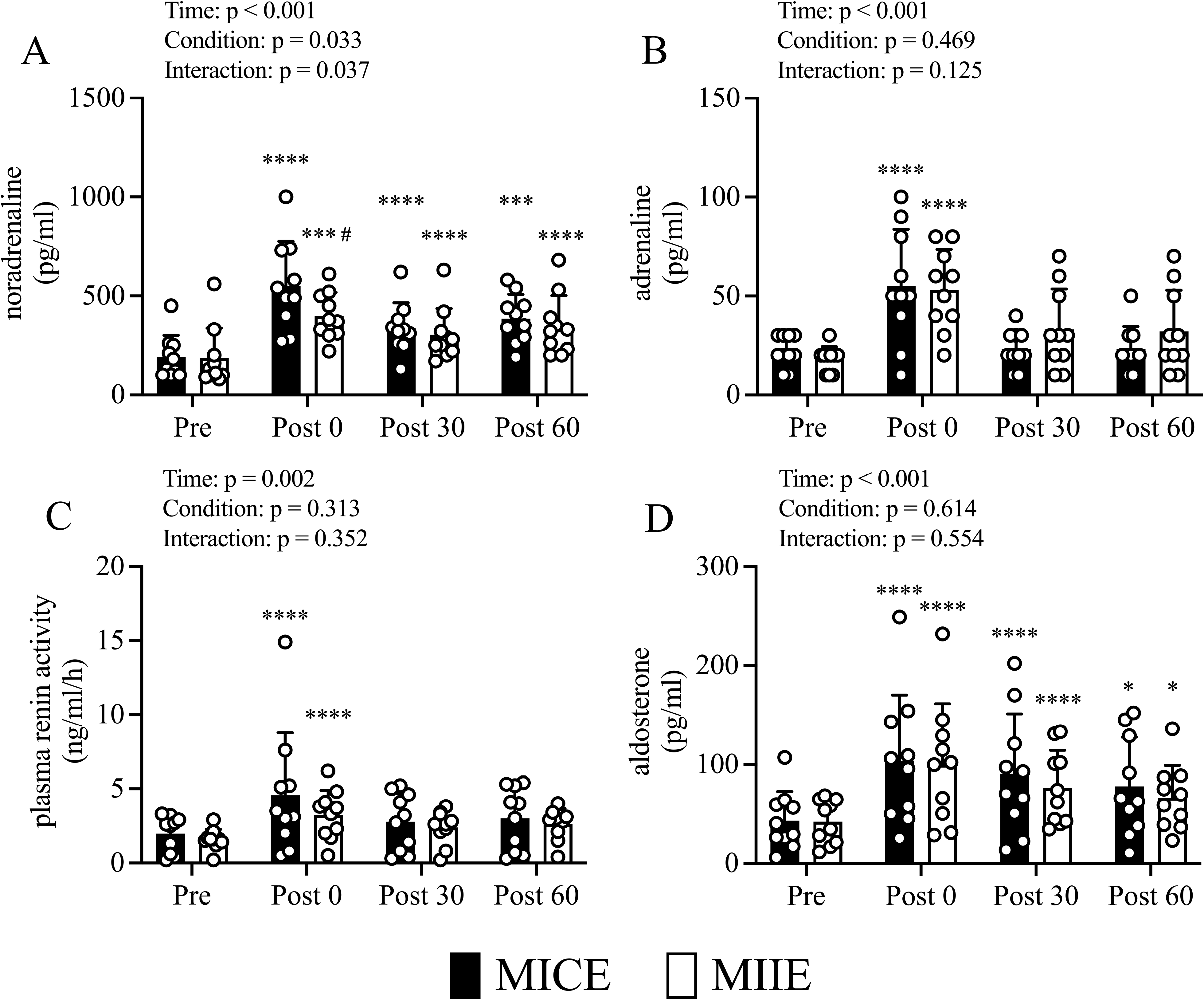
Changes in noradrenaline (**A**), adrenaline (**B**), plasma renin activity (**C**), aldosterone (**D**) from before exercise (*Pre*) to immediately after exercise (*Post 0*) to 30 min post-exercise (*Post 30*) to 60 min post-exercise (*Post 60*) in MICE (filled bar, n = 10) and MIIE (open bar, n = 10). Data are the mean ± standard deviation. * p < 0.05, ** p < 0.01, *** p < 0.005, **** p < 0.001 vs. *Pre*. ♯ p < 0.05 vs. MICE. Open circles represent individual data. Multiple pairwise comparisons were corrected using the Bonferroni method

There was no condition-by-time interaction effect for changes in adrenaline (Fig. 5B), PRA (Fig. 5C), or aldosterone (Fig. 5D) levels; however, there was a time effect for adrenaline (p < 0.001), PRA (p = 0.002) and aldosterone (p < 0.001) levels. Compared to *Pre*, adrenaline (p < 0.001) and PRA (p < 0.001) increased significantly at *Post 0* and showed no significant changes at *Post 30* and *Post 60* in either condition. Aldosterone levels were significantly higher at *Post 0* than at *Pre* (p < 0.001) and remained significantly higher at *Post 30* (p < 0.001) and *Post 60* (p = 0.017) under both conditions.

### Physiological variables following MICE and MIIE conditions

The variations in physiological parameters before and immediately after exercise and during the recovery period under both conditions (Table 2). There were no condition-by-time interaction effects; however, there was a time effect (p < 0.001) on heart rate. A condition-by-time interaction effect was observed regarding systolic blood pressure (p = 0.011). Systolic blood pressure before exercise did not differ between conditions, and systolic blood pressure exhibited a significant increase after exercise compared to before exercise in both conditions (p < 0.01, p < 0.01, respectively). However, the systolic blood pressure after exercise was lower in the MIIE condition than in the MICE condition (p = 0.046). There were no condition-by-time interaction, condition, or time effects on the changes in diastolic blood pressure and blood lactate concentrations.

**Table 2.** Comparison of physiological variables before and after MICE and MIIE, and recovery phase.

|  | MICE (n = 10) |  | MIIE (n = 10) |  | Interaction<br>effect | Time<br>effect | Condition<br>effect |
| --- | --- | --- | --- | --- | --- | --- | --- |
|  | before Ex | after Ex | before Ex | after Ex |  |  |  |
| Heart rate, bpm | 67 ± 10 | 100 ± 14 | 66 ± 10 | 94 ± 12 | p = 0.111 | p < 0.001 | p = 0.042 |
| Systolic blood pressure, mmHg | 107 ± 10 | 146 ± 15** | 108 ± 8 | 139 ± 16**, # | p = 0.011 | p < 0.001 | p = 0.117 |
| Diastolic blood pressure, mmHg | 76 ± 10 | 76 ± 13 | 75 ± 12 | 78 ± 13 | p = 0.506 | p = 0.767 | p = 0.836 |
| Blood lactate concentration, mmol/L | 1.5 ± 0.3 | 1.9 ± 0.4 | 1.6 ± 0.3 | 1.8 ± 0.3 | p = 0.399 | p = 0.06 | p = 0.693 |

Data are expressed as the mean SD.
**MICE** moderate-intensity continuous exercise, **MIIE** moderate-intensity intermittent exercise.
\* p < 0.05, \*\* p < 0.01 vs. before Ex. # p < 0.05, ## p < 0.01 vs. MICE

## Discussion

We evaluated the effects of intermittent and continuous exercise on renal haemodynamic and functional parameters during moderate-intensity exercise using ultrasound echo. The effects of MIIE and MICE exercise on AKI biomarkers were also assessed. Contrary to our hypothesis, MIIE had a similar effect on renal haemodynamics and function, and AKI biomarkers compared to MICE. MIIE and MICE maintained the RBF, eGFR, and FF levels and did not induce changes in AKI biomarkers. Notably, noradrenaline concentrations immediately after exercise were significantly lower under MIIE conditions than under MICE conditions.

The intensity of exercise may be a crucial factor affecting renal haemodynamics and AKI biomarkers. A moderate intensity is recommended as an intensity for individuals with CKD (Liguori et al. 2021). Many previous studies have reported that moderate-intensity exercise has beneficial effects in non-dialysis patients with CKD (Howden et al. 2012; Vanden Wyngaert et al. 2018; Watson et al. 2018; Kirkman et al. 2019; Thompson et al. 2019); however, there are few reports on the effects of moderate-intensity exercise on renal function (Greenwood et al. 2015; Santana et al. 2017; Bongers et al. 2017; Kawakami et al. 2022). According to previous studies (Wołyniec et al. 2020; Kosaki et al. 2020; Juett et al. 2021), a significant increase in AKI biomarkers was observed after high-intensity exercise, suggesting the possibility that glomerular abnormalities, tubular hypoxia, and tubulointerstitial damage are caused by exercise. Moreover, renal haemodynamics are strongly influenced by exercise intensity; RBF decreases in an intensity-dependent manner with exercise through the distribution of blood flow (Kawakami et al. 2018; Kotoku et al. 2019). However, these studies revealed that RBF does not decrease until moderate intensity and then decreases in an intensity-dependent manner at higher intensities. Therefore, participants who performed exercise at LT intensity did not have a decrease in RBF as moderate intensity, and there were no significant changes in AKI biomarkers following moderate-intensity exercise in the current study.

It is crucial for patients with CKD to exercise regularly. However, some patients with CKD may not be able to exercise continuously because of reduced cardiopulmonary function and skeletal muscle mass; therefore, the ACSM guidelines state that they should perform moderate-intensity exercise at intervals (Liguori et al. 2021). Exercise duration may be another crucial factor affecting central and peripheral circulation. A previous study (Seeger et al. 2015) examining the acute effects of intermittent exercise (IE) or continuous exercise (CE) on the vascular endothelium found that a single bout of IE, but not of CE, prevented endothelial ischaemia-reperfusion injury. Moreover, aerobic interval training promotes larger size and thickness of the renal tubule and improves renal functionality (Martínez et al. 2019). More recently, Meric et al. reported that a single bout of MIIE seems to have an effect on vascular endothelial function similar to that of MICE (Meric et al. 2022). Therefore, MIIE has been shown to have beneficial effects, and is recommended as part of a well-balanced exercise program. To the best of our knowledge, this is the first study to compare the acute effects of MIIE and MICE on renal haemodynamics.

A decrease in RBF due to exercise increases the glomerular permeability and enhances the filtration fraction (Poortmans et al. 1996; Svarstad et al. 2002), resulting in a decrease in the glomerular filtration rate and transient proteinuria (Grimby 1965; Clorius et al. 1996). Furthermore, a decrease in RBF with exercise induces a transient decrease in the peritubular capillary blood flow, which may lead to acute adverse effects on tubular conditions (Kosaki et al. 2020). Therefore, it is important to explore the effects of exercise on renal haemodynamics. We compared the effects of MIIE and MICE on renal haemodynamic parameters using ultrasound echo and showed that MIIE and MICE did not cause a significant change in BFV or CSA and consequently did not decrease RBF. Therefore, these findings demonstrate that MIIE and MICE have comparable effects on renal haemodynamic parameters. We also examined the effects of MIIE and MICE on catecholamine levels and RAS parameters to explore the mechanisms underlying renal haemodynamic regulation. Our detailed analysis revealed that adrenaline, PRA, and aldosterone had similar patterns under MIIE and MICE conditions. However, MIIE had a different effect on noradrenaline levels than MICE. A significant increase in noradrenaline concentration was observed immediately after MIIE and MICE. Moreover, the noradrenaline concentrations immediately after MIIE were significantly lower than those immediately after MICE. These findings demonstrate that IE suppressed the increase in noradrenaline concentration compared to CE during moderate-intensity exercise, suggesting that MIIE does not stimulate the sympathetic nervous system compared to MICE and is recommended to patients with CKD who have the sympathetic overactivity.

A few studies have examined the acute effects of IE and CE on the biomarkers of kidney injury. A recent study investigating the effect of manipulating hydration status during high-intensity intermittent running on AKI biomarkers suggested that hypohydration produced by high-intensity intermittent running increased renal injury compared to when euhydration was maintained (Juett et al. 2021). Houck et al. (Houck et al. 2022) examined the effects of MIIE and MICE on AKI biomarkers and renal function in hot environments. They revealed that MIIE and MICE in hot environments caused an increase in the biomarkers of kidney injury. Furthermore, we recently examined the acute effects of HIIE and MICE on AKI biomarkers in temperature-controlled conditions and demonstrated that glomerular or proximal tubular injury was not induced following a single bout of HIIE or MICE (Kawakami et al. 2024). Exercise-induced increases in AKI biomarkers appear to be dependent on the intensity or duration of exercise and the surrounding environment and have been shown to influence AKI biomarkers, which is primarily attributed to a greater magnitude of hyperthermia and hypovolaemia (Schlader et al. 2017; Chapman et al. 2020). In this study, we compared the acute effects of MIIE and MICE on AKI biomarkers under temperature-controlled conditions and demonstrated that neither MIIE nor MICE impaired the glomeruli or tubules. The assessment of urinary AKI biomarkers is likely influenced by changes in hydration status, and urinary biomarkers have been corrected for urinary osmolality (Juett et al. 2021; Houck et al. 2022). However, urinary osmolality was not measured in this study, and the effect of exercise on AKI biomarkers could not be examined in the context of hydration status. Therefore, further studies are needed to examine changes in urinary biomarkers corrected for urinary osmolality for hydration status.

Our study had some limitations. First, one of the main limitations was its relatively small sample size, which should be considered when interpreting the results. Secondly, we included middle-aged males (37 ± 8 years) with relatively preserved renal function (eGFR ≥ 60 ml/min/1.73 m^2^) in this study. The renal haemodynamic response to exercise may vary depending on the severity of the renal functional impairment. Therefore, similar studies using non-dialysis patients with CKD (eGFR < 60 ml/min/1.73 m^2^) should be performed in the future for clinical application.

In the current study, we investigated whether the effects of MIIE on renal haemodynamic and functional parameters as well as AKI biomarkers were different from those of MICE. Our findings demonstrated that there were no differences in renal haemodynamic responses to MIIE and MICE and that eGFR and FF exhibited a similar pattern under both conditions. MIIE and MICE did not alter the uL-FABP and uKIM-1 levels, indicating that neither MIIE nor MICE affected the glomeruli or tubules. Furthermore, our study provides novel evidence that moderate-intensity exercise with intervals suppresses the increase in noradrenaline concentration, suggesting that MIIE does not stimulate the sympathetic nervous system as compared to MICE. These findings provide additional data for designing effective exercise programs to prevent renal function decline, which could significantly contribute to the development of appropriate medical care.

To date, several studies have demonstrated the benefits of exercise in patients with CKD (Greenwood et al. 2015; Kirkman et al. 2019; Beetham et al. 2022), and exercise therapy may be an effective non-pharmacologic strategy. However, ideal exercise training programs for individuals with CKD have not yet been fully developed (Liguori et al. 2021). Therefore, it is important to accurately understand the effects of exercise conditions on kidneys to identify effective exercise conditions that minimise their burden. Our future perspective is to explore the effects of physical exercise on the kidney under different exercise conditions and to examine whether physical exercise can enhance or reversibly restore renal function. Ultrasound echo was used to examine the acute effects of MIIE or MICE on detailed renal haemodynamics, which minimised the burden on the participants. Therefore, ultrasound echoes may be clinically useful to noninvasively monitor renal haemodynamic changes following exercise.

## Conclusions

To our knowledge, this is the first study to investigate the effects of MIIE and MICE on renal haemodynamic parameters using ultrasound echo. Our findings demonstrated that there were no differences in the renal haemodynamic responses to MIIE and MICE. It also shows that MIIE and MICE do not alter urinary L-FABP and KIM-1 levels, indicating that neither condition affects the glomeruli or tubules. These findings suggest that MIIE has a similar effect on renal haemodynamics and function, and AKI biomarkers as MICE.

## Abbreviations

(s) Cr: (Serum) creatinine
(s) Cys: (Serum) cystatin C
(u) Alb: (Urinary) albumin
(u) Cr: (Urinary) creatinine
(u) L-FABP: (Urinary) liver-type fatty acid-binding protein
(u) uNAG: (Urinary) N-acetyl-beta-d-glucosaminidase
(u) uKIM-1: (Urinary) kidney injury molecule 1
(u) uNGAL: (Urinary) neutrophil gelatinase-associated lipocalin ACSM American College Sports Medicine
AKI: Acute kidney injury BFV Blood flow velocity BMI Body mass index
CKD: Chronic kidney disease CSA Cross sectional area
eGFR: estimated glomerular filtration rate ELISA Enzyme-linked immunosorbent assay FF Filtration fraction
MICE: Moderate intensity continuous exercise MIIE Moderate intensity intermittent exercise RAS Renin angiotensin system
RBF: Renal blood flow LT Lactate threshold
PRA: Plasma renin activity

## Acknowledgements

The authors thank the volunteers who participated in this study and acknowledge the contribution of the staff at Fukuoka University, who heled with the recruitment of participants.

## Author Contributions

S. Kawakami, T.Y., and R.M. conceived and designed research; S. Kawakami, T.Y., S.K., and A.I. performed experiments; S. Kawakami analyzed data; S. Kawakami, T.Y., and R.M. interpreted results of experiments; S. Kawakami prepared figures; S. Kawakami drafted manuscript; S. Kawakami, T.Y., S.K., A.I., K.F., T.M., S.N., K.M., Y.U., Y.H., and R.M. edited and revised manuscript; and all authors approved final version of manuscript.

## Data availability statement

The data that support the findings of the present study are available from the corresponding author upon reasonable request.

## Statements and Declarations

### Ethical approval

The study protocol (22-06-M1), which complies with the Declaration of Helsinki and the principles of Good Clinical Practice, was approved by the Ethics Committee of Fukuoka University. Participants in this study submitted signed informed consent after having all potential risks and procedures fully explained to them.

